# Prevalence of the photic sneeze reflex: A systematic review and meta-analysis

**DOI:** 10.64898/2026.08.10.26359569

**Authors:** Josef Trinkl, Stephan Munkwitz, Lucien Bickerstaff, Taisuke Eto, Manuel Spitschan

## Abstract

The photic sneeze reflex (PSR), in which exposure to bright light triggers sneezing, is widely recognised but inconsistently defined and measured. We conducted the first systematic review and meta-analysis of PSR prevalence to synthesise the epidemiological evidence, assess methodological quality, and identify priorities for future research. We included 18 articles comprising 31 study groups and extracted prevalence estimates, study characteristics, ascertainment methods, and epidemiological information. Fifteen eligible study groups classified as healthy were included in the primary meta-analysis. The pooled prevalence was 22% (95% CI, 15%-29%), with extreme between-study heterogeneity (I² = 99.3%). Reported prevalence estimates and associations with participant characteristics varied widely, and nearly all studies were judged to be at high risk of bias. The pooled estimate should therefore be interpreted as a descriptive summary of the available evidence rather than a precise estimate of population prevalence. Future studies require a standardised operational definition, representative sampling, transparent reporting, and reproducible methods for assessing light-triggered sneezing.

## Introduction

The photic sneeze reflex (PSR) is a recognisable human response in which sudden exposure to bright light, typically when moving from a darker environment into a well-illuminated one, elicits sneezing. The phenomenon has been described as the photic sneeze reflex, photic sneeze syndrome, sun sneezing, ACHOO (autosomal dominant compulsive helio-ophthalmic outburst) syndrome, and light-induced sneezing. Despite its everyday recognisability, the construct remains poorly standardised. Trigger conditions, latency, frequency, and number of sneezes vary between individuals and populations, although response patterns may be relatively stable within an individual (Semes et al., 1995). The absence of a shared operational definition complicates comparisons across studies.

Reported PSR prevalence estimates range from very low values to more than half of the sampled population. This variation may reflect biological differences, but it also arises from substantial methodological heterogeneity. Studies differ in their populations, recruitment settings, sampling procedures, demographic coverage, and treatment of clinical and ostensibly healthy groups. Ascertainment methods range from single self-report questions and questionnaires to clinical observation and experimental light provocation, with inconsistent wording and exposure conditions. Consequently, the prevalence of PSR is not only biologically uncertain but also methodologically difficult to estimate and interpret.

PSR is relevant to sensory integration and reflex physiology because it may reflect interactions between visual input and neural pathways involved in sneezing. Proposed mechanisms include cross-talk between visual and trigeminal pathways, general parasympathetic hypersensitivity, and parasympathetic generalisation through co-activation of closely situated nerve branches (Everett, 1964). These accounts remain plausible but unconfirmed, and no conclusive mechanistic model has been established.

Familial aggregation (Beckman and Nordenson, 1983; Everett, 1964; Forrester, 1985; Morris, 1987)and genome-wide association studies (Eriksson et al., 2010; Sasayama et al., 2018; Wang et al., 2019) suggest a heritable contribution to PSR. Understanding the conditions under which PSR occurs may also have practical relevance when unexpected sneezing could create risk, including during ocular procedures (Morley et al., 2010; Wessels et al., 1999). However, proposed mechanisms, inheritance patterns, and associations with participant characteristics remain unresolved.

No systematic review has previously synthesised the prevalence evidence for PSR or appraised the methodological quality of this literature. We therefore aimed to identify and summarise studies reporting PSR prevalence, extract and classify epidemiological information, assess risk of bias, estimate pooled prevalence in eligible healthy study groups, and define priorities for more standardised future research.

## Methods

### Study design and reporting framework

This systematic review and meta-analysis of PSR prevalence estimates was conducted and reported in accordance with the PRISMA 2020 statement (Page et al., 2021). No review protocol was registered.

### Eligibility criteria

Studies were eligible if they reported a PSR prevalence estimate or sufficient data to calculate one. We applied no restrictions by study design, population, or language. Eligible evidence included population surveys, clinical and laboratory studies, family studies, and reports in which PSR prevalence was recorded incidentally. Study groups preselected for photic sneezing were retained for the descriptive synthesis but excluded from pooled prevalence estimation.

### Information sources and search strategy

The principal information source was the bibliography compiled previously (Trinkl et al., 2025), which contained 167 records relating to PSR. The bibliography was assembled using MEDLINE (PubMed), Google Books, and Google searches with the terms “photic sneeze”, “sun sneeze*”, “bright light” AND “sneeze”, and variants in English, German, and French. Forward and backward citation searches in Google Scholar were used to supplement the bibliography, which was updated on 23 May 2021 and 31 January 2024 after the initial search on 9 November 2020. An additional unstructured Web of Science search on 10 December 2024 identified no further eligible articles. The bibliography is available at https://github.com/tscnlab/TrinklEtAl_ExpBrainRes_2025.

To update the existing reference library, we conducted a de novo MEDLINE search through PubMed using Medical Subject Headings (MeSH) and free-text terms. The complete Boolean search was: ((”Photic sneeze reflex”[MeSH] OR “photic sneeze”[tiab] OR “sun sneeze”[tiab] OR “ACHOO syndrome”[tiab] OR “light-induced sneeze”[tiab] OR “sneezing and light”[tiab] OR “sneeze reflex”[tiab] OR “sneezing response”[tiab])

AND

(”Prevalence”[MeSH] OR “epidemiology”[MeSH] OR prevalence[tiab] OR incidence[tiab] OR frequency[tiab] OR proportion[tiab] OR rate[tiab] OR distribution[tiab] OR epidemiologic study[tiab] OR population-based study[tiab] OR cross-sectional study[tiab] OR observational study[tiab] OR cohort study[tiab] OR case-control study[tiab] OR public health surveillance[tiab] OR health survey[tiab] OR national survey[tiab] OR risk factors[tiab] OR demographic factors[tiab] OR genetic predisposition[tiab]))

The final PubMed search was run on 28 June 2026 and returned 18 records.

### Study selection

In total, 180 records were screened by title, abstract, and, where appropriate, full text. Eighteen studies reporting PSR prevalence or data from which prevalence could be calculated were included. MS performed the initial screening and selection, and JT subsequently assessed the included studies and extracted the data. Duplicate records and reports without usable human PSR prevalence data were excluded. The study-selection process is summarised in the PRISMA flow diagram (**Figure 1**).

**Figure 1.**
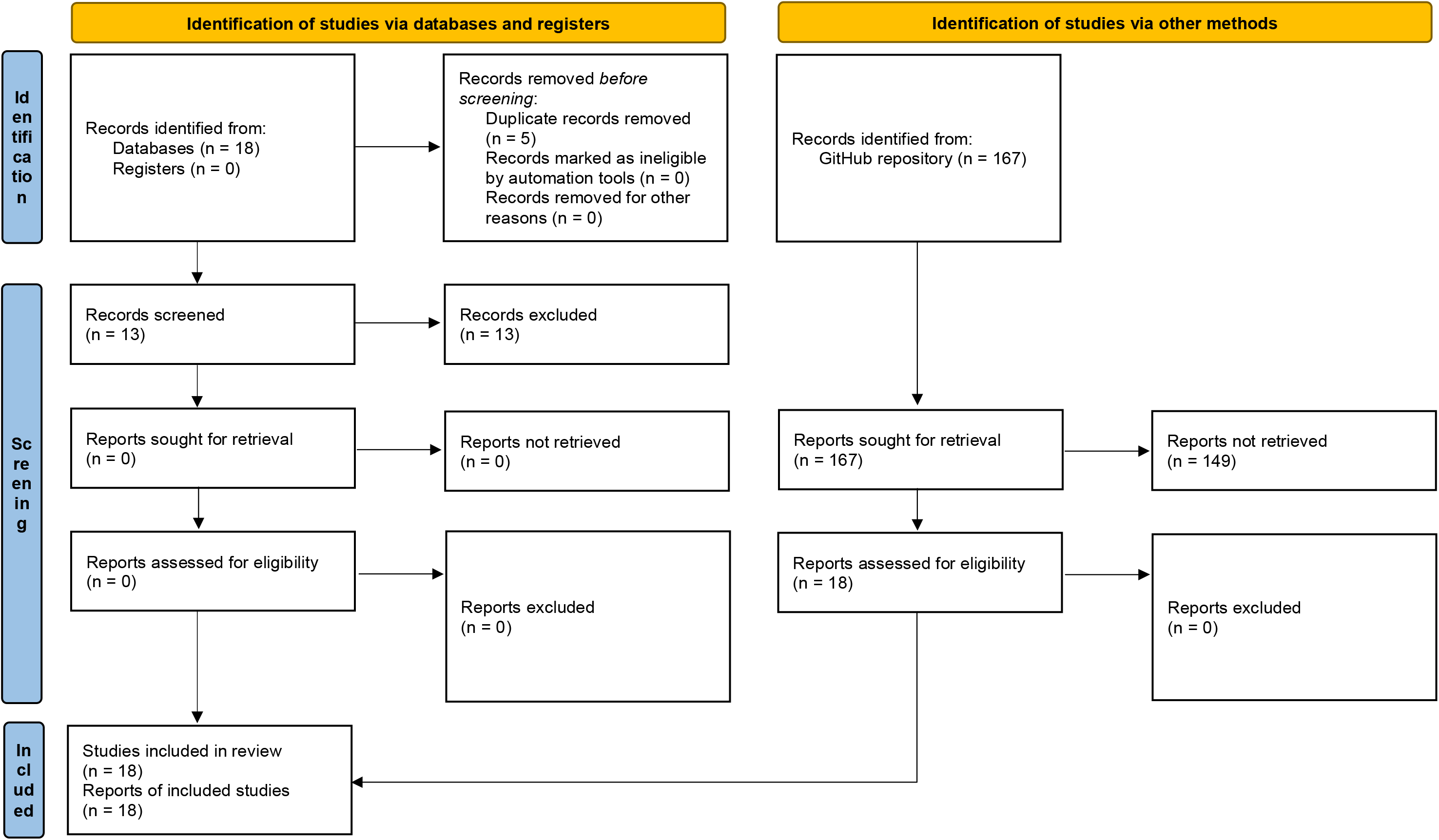
PRISMA 2020 flow diagram showing the study-selection process (adapted from Page et al., 2021).

**Figure 2.**
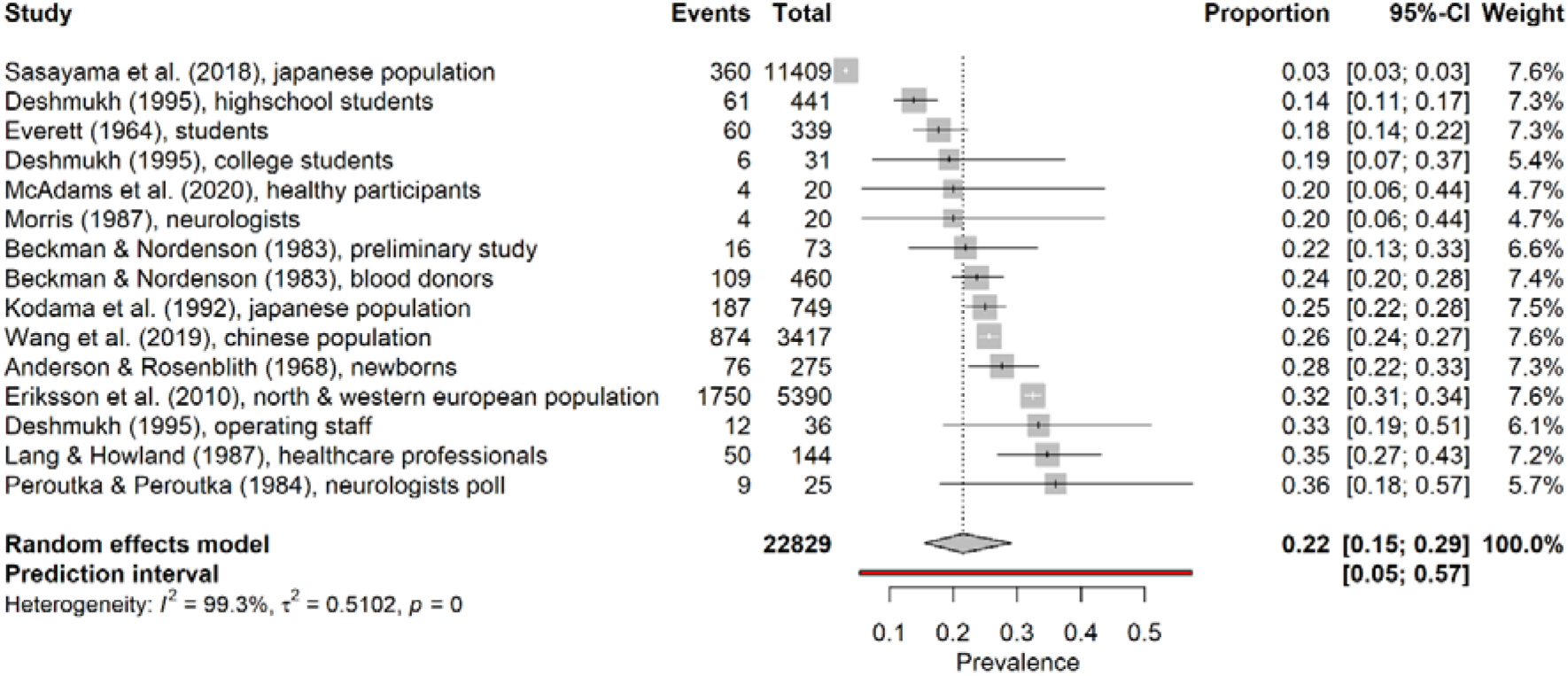
Forest plot of PSR prevalence in the 15 eligible healthy study groups.

**Figure 3.**
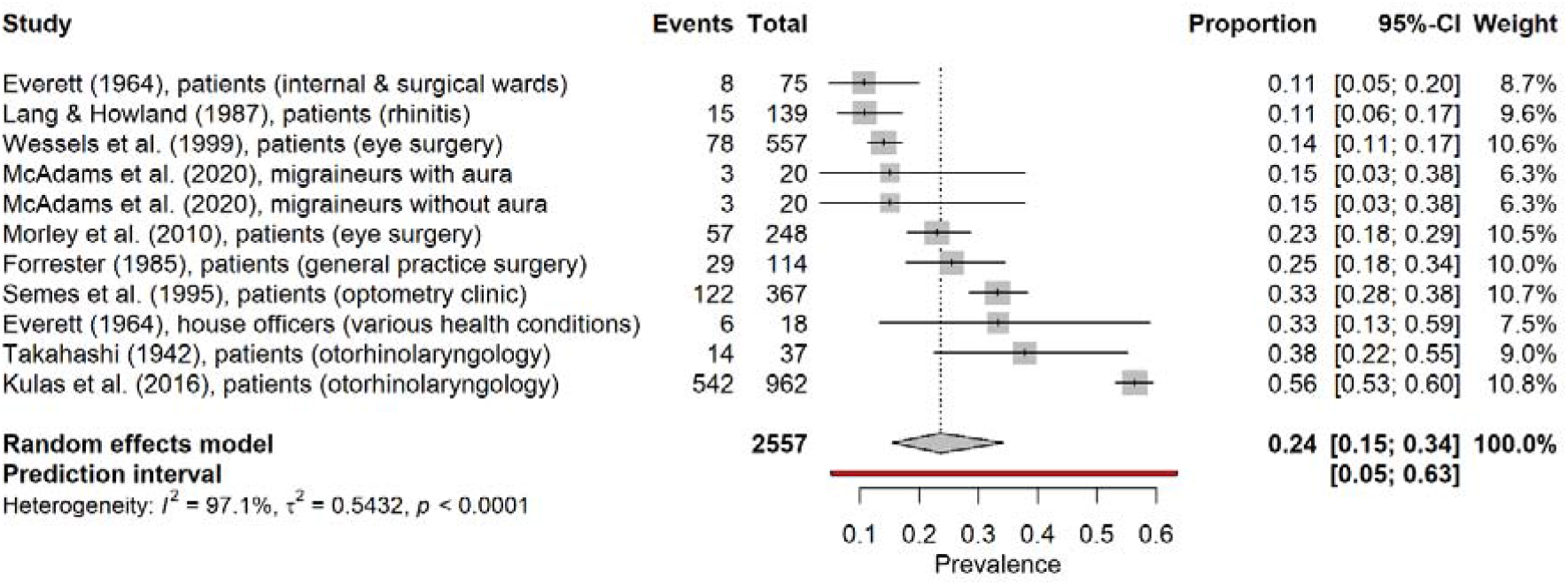
Forest plot of PSR prevalence in study groups classified as patient or clinical samples.

### Data extraction

For each eligible study, we extracted bibliographic information, study design, sample size, recruitment method, country and setting, participant demographics, occupation, and reported health status. We also recorded inclusion and exclusion criteria, the operational definition of PSR, the exact ascertainment question where available, the method used to identify photic sneezers, the light source and exposure conditions, potential confounders, and the numbers of participants with and without PSR. When a report contained multiple populations or subgroups, these were extracted separately to support consistent classification and analysis.

### Classification of study populations and subgroups

Study groups were classified according to their reported health status as healthy or patient/clinical samples. Groups preselected because participants or families had PSR were described narratively but excluded from pooled prevalence estimates. The primary quantitative synthesis included eligible healthy study groups; patient and clinical groups were analysed separately as an exploratory comparison.

### Risk-of-bias assessment

Risk of bias was assessed using the Joanna Briggs Institute Critical Appraisal Checklist for Prevalence Studies (Joanna Briggs Institute, 2020), with the appraisal focused specifically on the validity of each reported PSR prevalence estimate. Two authors independently assessed each of the 31 study groups (JT with either SM or TE). Initial agreement was reached for 175 of 279 item-level judgements (62.7%). Differences arose mainly from the interpretation of items Q5 (targeting coverage bias) and Q7 (standardisation of measurement). The assessors agreed on operational rules for these items and resolved all remaining disagreements by consensus with reference to the JBI guidance (Munn et al., 2015). For Q1, the sampling frame was judged in relation to the PSR prevalence question rather than the original purpose of the study. For Q3, a required sample size of 289 was calculated using an expected prevalence of 0.25, a 95% confidence level, and precision of 0.05. Overall risk-of-bias judgements were based on the number and likely influence of item-level concerns on the prevalence estimate.

### Data synthesis

Study characteristics, ascertainment methods, prevalence estimates, reported associations, and methodological limitations were synthesised narratively. Individual studies were summarised to retain relevant contextual information that could not be represented in the quantitative analysis.

### Statistical analysis

Random-effects meta-analyses of proportions were conducted in R (version 4.5.1) using RStudio (version 2025.09.1+401) and the metaprop function of the meta package (version 8.2.1). (Balduzzi et al., 2019). For each study group, prevalence was calculated as the number of participants classified as having PSR divided by the total sample size. The Freeman-Tukey double-arcsine transformation was used to stabilise variances for proportions near the boundaries, and pooled estimates were back-transformed to the original proportion scale. Between-study heterogeneity was quantified using I² and τ². The primary analysis included eligible healthy study groups; patient and clinical groups were pooled separately for descriptive comparison.

### Data and code availability

The PSR bibliography is available at https://github.com/tscnlab/TrinklEtAl_ExpBrainRes_2025 (Trinkl et al., 2025). The study-level extraction sheet and item-level JBI risk-of-bias assessments are provided in the supplementary material. Analysis code and figure-generation materials should be deposited with the final submission.

## Results

Eighteen studies reported PSR prevalence or sufficient data to calculate it. Seven studies contributed more than one distinct population, yielding 31 study groups in total. Twenty groups were classified as healthy and 11 as patient or clinical samples. (**Table 1**) Five healthy groups were preselected for PSR and were therefore excluded from quantitative synthesis, leaving 15 eligible healthy groups for the primary meta-analysis. Patient and clinical groups were analysed separately as an exploratory comparison.

**Table 1.** Characteristics of the included studies and study groups. m, male; f, female.

| Study group | Country | Population | Health status category | Sample size | Age | Sex | PSR definition | Diagnostic method | PSR question | Light stimulus | Prevalence proportion |
| --- | --- | --- | --- | --- | --- | --- | --- | --- | --- | --- | --- |
| Sasayama et al. (2018) | Japan | Japanese population | Healthy | 11409 | Mean age: Sneezers: 46.3 years<br>Non-sneezers: 50.0 years | sneezers: 176 m, 184 f<br>non-sneezers: 5849 m, 5200 f | tendency to sneeze when the eye is exposed to bright light | questionnaire | individuals who checked "light sneeze reflex" in a list of medical conditions/diseases in online survey were treated as PSR cases. | bright light | 0.03 |
| Everett (1964) house officers | USA | House officers | Patient | 18 | Adults | 6 m, 12 unclear | tendency to sneeze when suddenly exposed to bright light | self-report | not specified | bright light | 0.33 |
| Everett (1964) Patients |  | Medical and surgical patients | Patient | 75 | Adults | 49 m, 26 f |  | self-report |  |  | 0.11 |
| Everett (1964) Students |  | Medical students, college juniors & seniors | Healthy | 339 | Young adults | 193 m, 146 f |  | questionnaire |  |  | 0.18 |
| Lang & Howland (1987) Patients | USA | Rhinitis patients | Patient | 139 | 24 to 61 years | 5 m, 10 f | Sneezing upon entering sunlight; Tendency to sneeze one to 20 times in sunlight | self-report | not specified | sunlight | 0.11 |
| Lang & Howland (1987) healthcare professionals |  | Healthcare professionals | Healthy | 144 | not specified | unclear |  |  |  |  | 0.35 |
| Deshmukh (1995) college students | USA | College students | Healthy | 31 | Mean age: 20 years | unclear | reflex action when exposed to sudden bright light | self-report | not specified | bright light | 0.19 |
| Deshmukh (1995) operating staff |  | Operating room staff | Healthy | 36 | Mean age: 38 years |  |  |  |  |  | 0.33 |
| Deshmukh (1995) highschool students |  | High school students | Healthy | 441 | Mean age: 16 years |  |  |  |  |  | 0.14 |
| Wessels et al. (1999) | USA | Patients undergoing eye surgery | Patient | 557 | 16 to 97 years<br>Mean age: 69.9 years | 223 m, 334 f | sneezing is provoked by exposure to bright light | self-report | Have you ever noticed yourself sneezing when going into the sun or into bright light? | bright light | 0.14 |
| McAdams et al. (2020) healthy participants | USA | Control group | Healthy | 20 | Mean age: 31 years | 7 m, 13 f | sneezing in response to sunlight | self-report | subjects were asked if they "tend to sneeze when [they] step out of a dark room into bright sunlight" | bright sunlight | 0.20 |
| McAdams et al. (2020) migraineurs with aura |  | Migraineurs with aura | Patient | 20 | Mean age: 31 years | 1 m, 19 f |  |  |  |  | 0.15 |
| McAdams et al. (2020) migraineurs without aura |  | Migraineurs without aura | Patient | 20 | Mean age: 30 years | 3 m, 17 f |  |  |  |  | 0.15 |
| Morris (1987) neurologists | USA | Neurologist Grand Round | Healthy | 20 | not specified | 15 m, 5 f | sneezing on exposure to bright light | questionnaire | not specified | bright light | 0.20 |
| Morley et al. (2010) | UK | Patients undergoing eye surgery | Patient | 248 | 19 to 105 years | unclear | sneezing on exposure to bright sunlight | self-report | not specified | bright sunlight | 0.23 |
| Beckman & Nordenson (1983) blood donors | Sweden | Blood donors | Healthy | 460 | not specified | unclear | sudden exposure to strong light may induce sneezing | self-report | not specified | bright light | 0.24 |
| Beckman & Nordenson (1983) preliminary study |  | Families from a preliminary study | Healthy | 73 |  |  |  | not specified |  |  | 0.22 |
| Kodama et al. (1992) | Japan | Japanese population | Healthy | 749 | 10 to 69 years | 391 m, 358 f | light-sneeze reflex | questionnaire or interview | not specified | bright light | 0.25 |
| Kulas et al. (2016) | Germany | Patients in otorhinolaryngology outpatient center | Patient | 962 | not specified | 440 m, 534 f | sneezing in response to bright light, sunlight and artificial light | questionnaire | not specified | bright light | 0.56 |
| Forrester (1985) | UK | Patients at a general surgery practice | Patient | 114 | not specified | unclear | sneezing in response to bright light | unclear | not specified | bright light | 0.25 |
| Wang et al. (2019) | PR China | Chinese population | Healthy | 3417 | 16 to 70 years | 1700 m, 1717 f | uncontrollable reflexive sneezing in response to sudden exposure to bright light | self-report | "in most cases, when you enter from a dark or dim environment suddenly into a bright environment, would you: A. sneeze, B. be a little responsive but do not sneeze, and C. have no response" | bright light/environment | 0.26 |
| Anderson & Rosenblith (1968) | USA | Newborns | Healthy | 275 | Newborns | 137 m, 138 f | sneezing in response to bright light | clinical observation | not specified | reflection of a shiny bicycle bell or flashlight shining into the eyes | 0.28 |
| Eriksson et al. (2010) | Online | Northern European descent | Healthy | 5390 | not specified | unclear | tendency to sneeze when moving from relative darkness into bright light — most often sunlight | questionnaire | Do you have a tendency to sneeze when exposed to bright sunlight? | bright light; emphasis on sunlight | 0.32 |
| Semes et al. (1995) | USA | Patients receiving | Patient | 367 | 12 to 89 years | unclear | sneezing in response to | questionnaire | not specified | unclear. Some answers | 0.33 |
|  |  | eye examination |  |  | Mean age<br>sneezers:<br>45.8 years<br>non-sneezers:<br>44.0 years |  | bright light |  |  | refer to sunlight, but it<br>was also asked for<br>sneezing in response to<br>other bright light stimuli |  |
| Peroutka & Peroutka (1984)<br>neurologists poll | USA | Neurologists | Healthy | 25 | not specified | unclear | sneezing in response to<br>bright light | poll between<br>neurologists | not specified | bright light | 0.36 |
| Takahashi (1942) | Japan | Rhinitis patients at<br>outpatient clinic | Patient | 79 | 1 to 60 (2 cases<br>over 60) | 62 m, 17 f | sneezing in response to<br>bright light (Lichtniesen) | clinical<br>observation &<br>direct<br>interview | not specified | bright light | 0.38 |

Nearly all included studies were judged to be at high risk of bias (**Table 2**). The most frequent concerns involved the representativeness of the sampling frame and recruitment method (Q1 and Q2), sample coverage and response-rate management (Q5 and Q9), and the validity and consistency of PSR ascertainment (Q6 and Q7). Most studies used convenience or otherwise narrowly defined samples, provided incomplete demographic information, or did not report the exact wording used to identify PSR. Definitions also differed in whether they referred specifically to sunlight, included artificial light, or left the light source unspecified. Although several studies reported high response rates and analysed the identified samples appropriately, these strengths did not resolve the broader limitations in sampling and outcome measurement.

**Table 2.** Item-level risk-of-bias judgements using the JBI Critical Appraisal Checklist for Prevalence Studies, visualised in a colour-blind-friendly format adapted from robvis (McGuinness and Higgins, 2021).

|  | Risk of bias |  |  |  |  |  |  |  |  |  |
| --- | --- | --- | --- | --- | --- | --- | --- | --- | --- | --- |
|  | D1 | D2 | D3 | D4 | D5 | D6 | D7 | D8 | D9 | Overall |
| Sasayama et al. (2018) | X | X | + | X | + | X | + | X | + | X |
| Everett (1964), house officers | X | X | X | + | X | X | X | X | X | X |
| Everett (1964), patients | X | X | X | + | + | X | X | X | + | X |
| Everett (1964), students | X | X | + | + | + | X | + | X | + | X |
| Lang & Howland (1987), patients | X | X | X | X | X | X | X | X | - | X |
| Lang & Howland (1987), healthcare professionals | X | X | X | X | X | X | X | X | - | X |
| Deshmukh (1995), college students | X | X | X | X | + | X | X | X | + | X |
| Deshmukh (1995), operating staff | X | X | X | X | + | X | X | X | + | X |
| Deshmukh (1995), highschool students | X | X | + | X | + | X | X | X | + | X |
| Deshmukh (1995), case report 1 | X | X | X | X |  | X | X |  |  | X |
| Deshmukh (1995), case report 2 | X | X | X | X |  | X | X |  |  | X |
| Wessels et al. (1999) | X | X | + | + | + | X | + | + | + | X |
| McAdams et al. (2020), healthy participants | X | X | X | X | + | X | X | X | + | X |
| McAdams et al. (2020), migraineurs with aura | X | X | X | X | + | X | X | X | + | X |
| McAdams et al. (2020), migraineurs without aura | X | X | X | X | + | X | X | X | + | X |
| Morris (1987), case report 1 | X | X | X | X |  | X | X |  |  | X |
| Morris (1987), case report 2 | X | X | X | X |  | X | X |  |  | X |
| Morris (1987), neurologists | X | X | X | X | + | X | + | X | - | X |
| Morley et al. (2010) | X | X | X | + | X | X | X | X | X | X |
| Beckman & Nordenson (1983), blood donors | X | X | + | X | + | X | X | X | + | X |
| Beckman & Nordenson (1983), preliminary study | X | X | X | X | + | X | X | X | + | X |
| Kodama et al. (1992) | X | X | + | X | X | X | X | X | + | X |
| Kulas et al. (2016) | X | X | + | X | X | X | + | X | X | X |
| Forrester (1985) | X | X | X | X | + | X | X | X | + | X |
| Wang et al. (2019) | X | X | + | + | + | X | + | + | + | X |
| Anderson & Rosenblith (1968) | X | X | X | + | + | - | - | X | X | X |
| Eriksson et al. (2010) | + | + | + | + | + | + | + | X | + | + |
| Semes et al. (1995) | X | X | + | X | X | X | + | X | X | X |
| Peroutka & Peroutka (1984), neurologists poll | X | X | X | X | + | X | X | X | + | X |
| Peroutka & Peroutka (1984), case report | X | X | X | X |  | X | X |  |  | X |
| Takahashi (1942) | X | X | X | X | X | X | - | X | + | X |

D1: JBI Q1 D2: JBI Q2 D3: JBI Q3 D4: JBI Q4 D5: JBI Q5 D6: JBI Q6 D7: JBI Q7 D8: JBI Q8 D9: JBI Q9
Judgement X High + Unclear - Low • Not applicable

### Individual study summaries

#### Sneezing in response to light (Everett, 1964)

Everett investigated associations between PSR and heredity, sex, allergy, and nasal disorders in three samples: psychiatric house officers, 75 hospitalised medical and surgical patients, and students from three colleges. The student survey yielded 339 analysable responses. The study reported associations with sex and ethnicity but not with allergic rhinitis or other allergies, and proposed parasympathetic excitation as a possible common mechanism. The paper also discussed familial patterns, related reflexes, and possible practical implications.

#### The photic sneeze reflex in the human newborn: a preliminary report (Anderson and Rosenblith, 1968)

During a study of visual following in 275 newborns, the investigators presented a shiny but silent bicycle bell for 1-3 minutes and used a penlight when the bell elicited no response. Seventy-six infants were reported to have sneezed, including 36 who sneezed only during presentation of the bell. Among infants with documented sneeze counts, most sneezed once. Subgroup analyses considered sex and ethnicity, and a separate case described light-triggered sneezing in a newborn with trisomy 13.

#### Clinical and statistical observations on so-called vasomotor rhinitis (Takahashi, 1942)

Takahashi examined 79 patients with vasomotor rhinitis at the outpatient clinic of Kyoto Imperial University. Light-related sneezing was assessed in 37 patients, of whom 14 (37.8% ± 7.9%) sneezed after moving from indoors into direct sunlight. Some patients also responded to strong artificial light. On the basis of these observations, the author used the term “Lichtniesen” for light-induced sneezing.

#### Solar sneeze reflex (Lang and Howland, 1987)

Lang and Howland reported 15 patients with allergic or vasomotor rhinitis who sneezed on entering sunlight, corresponding to approximately 11% of the clinic population. After treatment of rhinitis, four patients no longer reported sunlight-triggered sneezing and three reported an attenuated response, although changes were not clearly related to rhinitis type. A separate survey of healthcare professionals at the same clinic identified PSR in 50 of 144 respondents (35%).

#### The photic sneeze reflex and ocular anaesthesia (Wessels et al., 1999)

Wessels et al. prospectively assessed PSR and involuntary sneezing during local anaesthesia in 557 patients undergoing ocular surgery. Before drug administration, participants were asked whether they had ever sneezed when entering sunlight or bright light; 78 (14%) responded affirmatively. Twenty-nine patients (5.2%) sneezed after anaesthesia. Sneezing was more frequent after periocular than retrobulbar injection, but the association between a history of PSR and procedure-related sneezing was not statistically significant. The study also found no significant difference in procedure-related sneezing between male and female patients.

#### Factors prompting sneezing during local anaesthetic injections to the eyelids (Morley et al., 2010)

Morley et al. analysed 314 procedures in 294 patients receiving intravenous sedation and local anaesthetic injections for oculoplastic surgery. A history of photic sneezing was reported by 23% of participants, and sneezing during or within 5 minutes of injection occurred in 51 procedures involving 47 patients. Procedure-related sneezing was associated with male sex, upper-eyelid infiltration, bilateral infiltration, deeper sedation, and midazolam use, whereas opioid derivatives were associated with lower risk. A history of photic sneezing was also associated with procedure-related sneezing (RR 2.6, 95% CI 1.6-4.0).

#### Selective amplification of ipRGC signals accounts for interictal photophobia in migraine (McAdams et al., 2020)

McAdams et al. studied visual discomfort and pupil responses in 20 control participants and 40 participants with migraine, divided according to the presence or absence of aura. As part of the assessment, participants were asked whether they tended to sneeze when moving from a dark room into bright light. Between 15% and 20% of each group responded affirmatively, with no significant between-group difference.

#### Individual differences in the sneezing reflex: an inherited physiological trait in humans? (Beckman and Nordenson, 1983)

Among 460 blood donors in Umeå, Sweden, 109 (24%) reported sneezing after sudden exposure to strong light. Women were more numerous among photic sneezers, but the association with sex was not statistically significant. The authors also reported a preliminary family study: in seven families with one photic-sneezing parent, 9 of 14 children reportedly had the trait, whereas no PSR was reported in 11 families in which neither parent had the trait.

#### Autosomal dominant transmission of the photic sneeze reflex (Peroutka and Peroutka, 1984)

A poll of 25 neurologists at Johns Hopkins Hospital identified PSR in 36%, although only 8% reported prior awareness of the phenomenon. The authors also described a family in which PSR was present in one author, his father, his brother, and subsequently his infant daughter, but not in his mother or wife. The family history was presented as evidence consistent with dominant transmission.

#### Sneezing on exposure to bright light as an inherited response (Forrester, 1985)

Forrester reported that 29 of 114 consecutive patients attending a general practice (25%) responded affirmatively when asked about sneezing in bright light. Six pedigrees were obtained. Across 15 couples in which one partner reported PSR, 17 of 37 children were also reported to have the trait.

#### ACHOO syndrome: prevalence and inheritance (Morris, 1987)

Morris presented two case reports with pedigrees interpreted as supporting autosomal dominant inheritance with variable expressivity. Some relatives also reportedly sneezed in response to eyebrow plucking or hair pulling. In an additional questionnaire administered at a meeting of neurologists, 4 of 20 respondents (20%) reported sneezing on exposure to bright light.

#### Questionnaire survey of sneezing induced by light stimulation in Tohoku (Kodama et al., 1992)

Kodama et al. surveyed 749 people aged from their teens to their sixties in workplaces, schools, and public settings in the Tohoku region of Japan. PSR was reported by 187 respondents (25%; 99 male and 88 female). Reported prevalence was highest among people in their twenties and somewhat lower in older age groups. Most photic sneezers reported a single sneeze at the moment of glare, although a small minority reported occasional multiple sneezes. Responses indicated substantial individual variation in triggering conditions, including sunlight, artificial light, and the intensity of the transition. Forty-three respondents reported a relative with the same response.

#### Sneezing in response to bright light: is it a cause of accidents? (Deshmukh, 1995)

Deshmukh combined two family case reports with surveys of college students, high-school students, and operating-room staff. Across the three surveys, 79 of 508 respondents (15.6%) reported sneezing on sudden exposure to bright light. The family reports were interpreted as consistent with dominant inheritance, and the article discussed circumstances in which an unexpected sneeze could present a safety risk.

#### The photic sneeze response: a descriptive report of a clinic population (Semes et al., 1995)

Semes et al. distributed questionnaires to 500 consecutive patients attending a university optometric clinic; 367 questionnaires were returned and 122 respondents (33.2%) identified as photic sneezers. PSR was more frequently reported by White than Black respondents, and an association with self-reported deviated nasal septum was also observed, although relevant subgroup information was incomplete. Most photic sneezers reported that sunlight triggered sneezing rarely or occasionally, and 107 of 118 respondents with available data reported three or fewer sneezes per episode.

#### Prevalence of the photo-induced sneezing reflex in a German clinical sample (Kulas et al., 2017)

Kulas et al. administered a standardised questionnaire to 962 patients attending an otorhinolaryngology outpatient clinic. The questionnaire assessed occurrence and frequency of photic sneezing, additional triggers, avoidance strategies, demographic characteristics, familial predisposition, medical history, and substance use. PSR was reported by 542 participants (56%), although 301 described only occasional sneezing; excluding these responses produced an estimate closer to those reported in other studies. Familial predisposition and nicotine consumption were associated with PSR, whereas other examined associations were not statistically significant.

#### Web-based, participant-driven studies of common genetic traits (Eriksson et al., 2010)

Eriksson et al. analysed 535,076 single-nucleotide polymorphisms in customers of a direct-to-consumer genetic testing company across 22 traits. For photic sneezing, 1,750 of 5,390 participants of Northern European ancestry (32.5%) answered yes to the question, “Do you have a tendency to sneeze when exposed to bright sunlight?” The analysis identified one genome-wide significant intergenic locus on 2q22.3 and a suggestive association near NR2F2 on chromosome 15.

#### Genome-wide association study of photic sneeze syndrome in a Japanese population (Sasayama et al., 2018)

Sasayama et al. analysed survey and genotype data from 11,409 participants in Japan. Participants who selected “light sneeze reflex” from a list of medical conditions were classified as photic sneezers, producing a prevalence estimate of 3.2%. The analysis supported previously reported loci and identified two additional suggestive associations in the Japanese sample. The ascertainment method, which treated PSR as one item within a list of conditions, differed substantially from the more direct questions used in several other studies.

#### Genome-wide association study of the photic sneeze reflex in a Chinese population (Wang et al., 2019)

Wang et al. analysed questionnaire and genotype data from 3,417 participants in China and reported a PSR prevalence of 25.6%. Prevalence was higher in male than female participants (30.1% vs 21.1%). Two independent genetic variants were associated with PSR. One locus on 2q22.3 had been reported previously, whereas a locus on 3p12.1 was newly identified in this analysis. Both associated variants were intergenic, and the 3p12.1 locus was located near CADM2.

### Synthesis of results

The 18 included studies had sample sizes ranging from 6 to 11,409 participants and comprised analytical (n = 6) and descriptive (n = 12) observational designs. Study populations included community and online samples, students, healthcare professionals, blood donors, newborns, family members, and patients recruited in clinical settings. Because several reports included more than one population, 31 distinct study groups were extracted: 20 classified as healthy and 11 as patient or clinical samples. Five healthy groups were excluded from quantitative synthesis because they were preselected for PSR, leaving 15 groups in the primary meta-analysis.

Across the 15 eligible healthy study groups, prevalence estimates ranged from 3% to 36%. The random-effects pooled prevalence was 22% (95% CI, 15%-29%), with extreme heterogeneity (I² = 99.3%; τ² = 0.5102). Across patient and clinical groups, prevalence estimates ranged from 11% to 56%, with an exploratory pooled estimate of 24% (95% CI, 15%-34%) and similarly high heterogeneity (I² = 97.1%; τ² = 0.5432). These pooled values therefore summarise highly heterogeneous evidence and should not be interpreted as precise population estimates.

Almost all studies ascertained PSR by self-report, usually through a questionnaire or a direct question about sneezing after exposure to sunlight or bright light. Response formats ranged from binary self-identification to graded frequency or intensity scales and open-ended answers. Only four studies reported the exact wording of the ascertainment question (Eriksson et al., 2010; McAdams et al., 2020; Wang et al., 2019; Wessels et al., 1999) and only one study directly observed sneezing during light exposure (Anderson and Rosenblith, 1968).

All but one study were judged to be at high risk of bias under the JBI appraisal. The principal concerns were non-representative or poorly described sampling methods, inconsistent definitions of PSR and its triggering light source, limited validity of outcome ascertainment, and incomplete or inappropriate statistical analysis.

Reported associations with sex, age, ancestry or ethnicity, and health status were inconsistent and were not examined using comparable methods across studies. Several family and genetic studies suggested a heritable contribution to PSR, but the available evidence did not permit robust estimation of inheritance patterns or subgroup differences.

## Discussion

This review provides the first systematic synthesis of PSR prevalence studies, the first pooled prevalence estimate, an explicit appraisal of operational definitions and methodological quality, and a proposed framework for more standardised future assessment. The pooled estimate of 22% across 15 eligible healthy study groups suggests that PSR is not rare, but its interpretation is constrained by extreme heterogeneity and the high risk of bias in nearly all studies. Differences in recruitment, population characteristics, ascertainment questions, light sources, and response definitions mean that studies may not have measured the same underlying construct. The exploratory analysis of patient and clinical samples showed a similar pooled estimate, but the constituent populations were diverse and do not define a single clinically meaningful target population. Reported associations with sex, age, ancestry or ethnicity, health status, and familial occurrence were inconsistent and should not be treated as established.

The review has several strengths. It draws together literature spanning multiple decades, languages, settings, and study designs; separates distinct study groups; and applies a structured prevalence-specific risk-of-bias appraisal. Its main limitation is the quality of the underlying evidence. Many studies were not designed primarily to estimate PSR prevalence, most relied on convenience samples and self-report, and almost all were at high risk of bias. The evidence base also overrepresents populations of European ancestry and provides insufficient information for reliable comparison across geographic or demographic groups. Accordingly, the pooled prevalence is best understood as a descriptive summary of the available literature rather than a general or global population estimate.

Future studies should use a shared operational definition and distinguish self-reported sunlight-triggered sneezing from responses observed during controlled light provocation. A minimum assessment should include a clearly worded screening question, graded measures of frequency and reproducibility, latency and sneeze count, and separate questions about sunlight and artificial light. Experimental studies should report the source, spectrum, intensity, geometry, duration, and temporal profile of light exposure, as well as pre-exposure conditions. Epidemiological studies require more representative sampling and fuller reporting of recruitment setting, ancestry or ethnicity, age, sex and gender, health status, relevant clinical conditions, medications, and other sneeze triggers. These improvements would allow future research to determine whether observed subgroup differences reflect biological variation or methodological artefact. To facilitate harmonisation across studies, we additionally propose a standardised PSR screening questionnaire incorporating these core elements. The questionnaire is intended as a practical minimum screening tool for self-reported photic sneeze responses and can be included as a supplementary assessment alongside study-specific measures.

### Proposed minimum recommendations for future PSR prevalence studies

**<u>Operational definition</u>**

- The photic sneeze reflex is a predisposition to sneeze following exposure to a bright light stimulus or a sudden increase in ambient light intensity.

**<u>Recommended screening question</u>**

- Do you sneeze when you are suddenly exposed to bright light, for example when stepping from a darker place into sunlight? Yes / No / Unsure / Light-induced nasal sensation without sneezing

**<u>Frequency, latency, multiplicity, and reproducibility</u>**

- How often do you sneeze when exposed to bright light? Always / Often / Sometimes / Rarely / Never
- What is the typical latency between light exposure and the first sneeze?
- How many sneezes typically occur during a light-triggered episode?
- What is the typical interval between successive sneezes?
- Has this response occurred consistently across your lifetime? Yes / No / Unsure
- When the same light transition is repeated, how often does it trigger sneezing? Always / Often / Sometimes / Rarely / Never

**<u>Sunlight and artificial light</u>**

- Does natural sunlight trigger sneezing? Yes / No / Unsure
- Does artificial light trigger sneezing? Yes / No / Unsure

**<u>Distinguish self-reported and experimentally provoked PSR</u>**

- State whether PSR was assessed by structured questionnaire or interview or observed during controlled light stimulation. Report the instrument, mode of administration, outcome definition, and criteria used to classify a participant as having PSR.

**<u>Report light exposure conditions and participant characteristics</u>**

- Report the light source; direct or diffuse illumination; continuous, pulsed, or flash exposure; spectral characteristics; illuminance at eye level; pre-exposure conditions; duration and rate of the dark-to-light transition; time of day; relevant environmental conditions; and the use of glasses, sunglasses, or contact lenses.
- Report participant age, sex and gender, ancestry or ethnicity, family history of PSR, allergic or non-allergic rhinitis, migraine, photophobia, relevant ophthalmological or neurological conditions, smoking and exposure to nasal irritants, medications that may affect sneezing, and other substances or stimuli that trigger sneezing.

## Conclusion

This systematic review identified a substantial but methodologically weak literature on PSR prevalence. The pooled estimate of 22% across eligible healthy study groups indicates that PSR is commonly reported, but extreme heterogeneity and pervasive risk of bias prevent precise inference about prevalence in any general or global population. The principal limitations are inconsistent definitions, non-representative sampling, and reliance on poorly documented self-report measures. A standardised operational definition, reproducible ascertainment methods, representative sampling, and transparent reporting are required before the epidemiology and determinants of PSR can be characterised reliably.

## Supplementary information and declarations

### Supplementary information

Data extraction form and item-level risk-of-bias assessments

## Authors’ contributions

Conceptualisation: MS. Data Curation: MS, JT, TE. Formal Analysis: JT, SM, TE. Funding Acquisition: MS. Investigation: JT, MS. Methodology: JT, MS. Project Administration: MS. Resources: –. Software: –. Supervision: MS. Validation: –. Visualisation: –. Writing – Original Draft Preparation: JT. Writing – Review & Editing: JT, SM, LB, TE, MS.

## Data availability

The study-level data, risk-of-bias assessments, analysis code, and figure-generation materials will be made available under a CC BY licence in the project repository [permanent repository URL to be added].

## Declarations

### Ethics approval and consent to participate

Not applicable.

### Consent for publication

Not applicable.

### Declaration of AI-assisted technologies

During preparation of this manuscript, the authors used ChatGPT to support language editing and the organisation of information. The authors subsequently reviewed and revised all content and take full responsibility for the final manuscript.

## Photic Sneeze Survey Questionnaire

Please complete the survey below. Thank you!

Thank you for agreeing to participate in this survey.

Please note: Some questions may seem repetitive. We include these to check consistency. Please answer all questions to the best of your knowledge and ability.

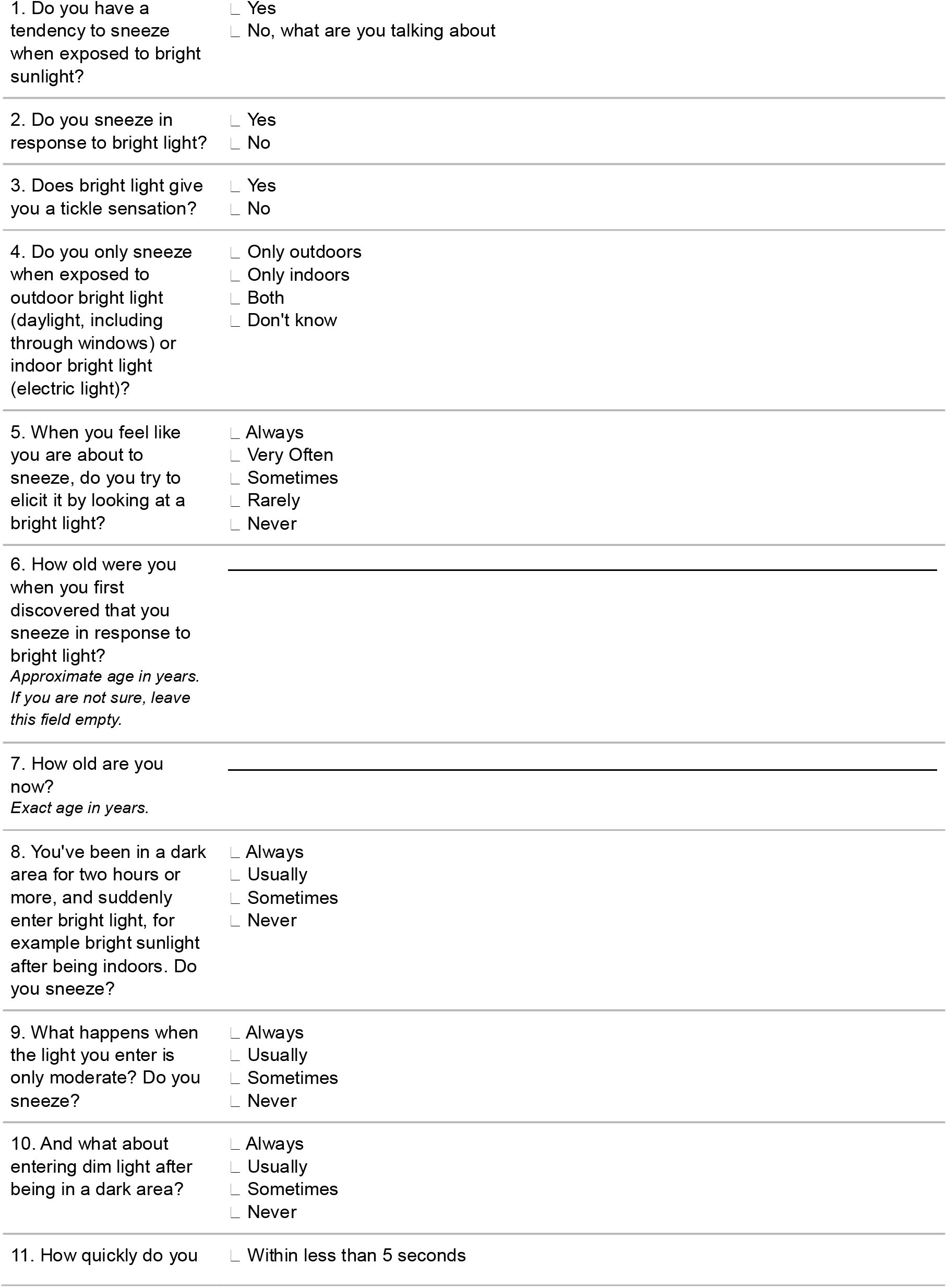
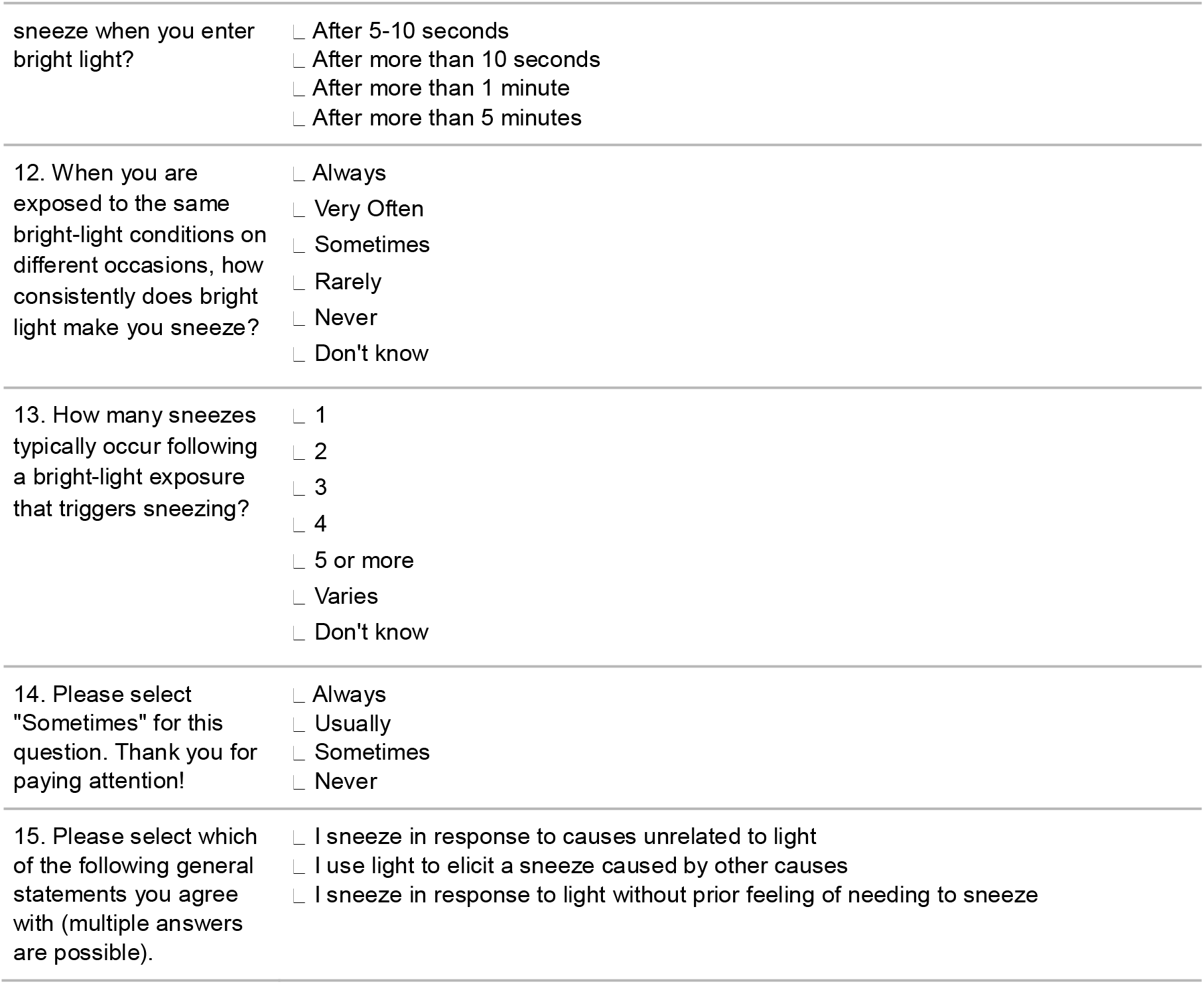

## Sneezing in daylight

Please select to what extent exposure to different levels of DAYLIGHT elicits a sneeze.

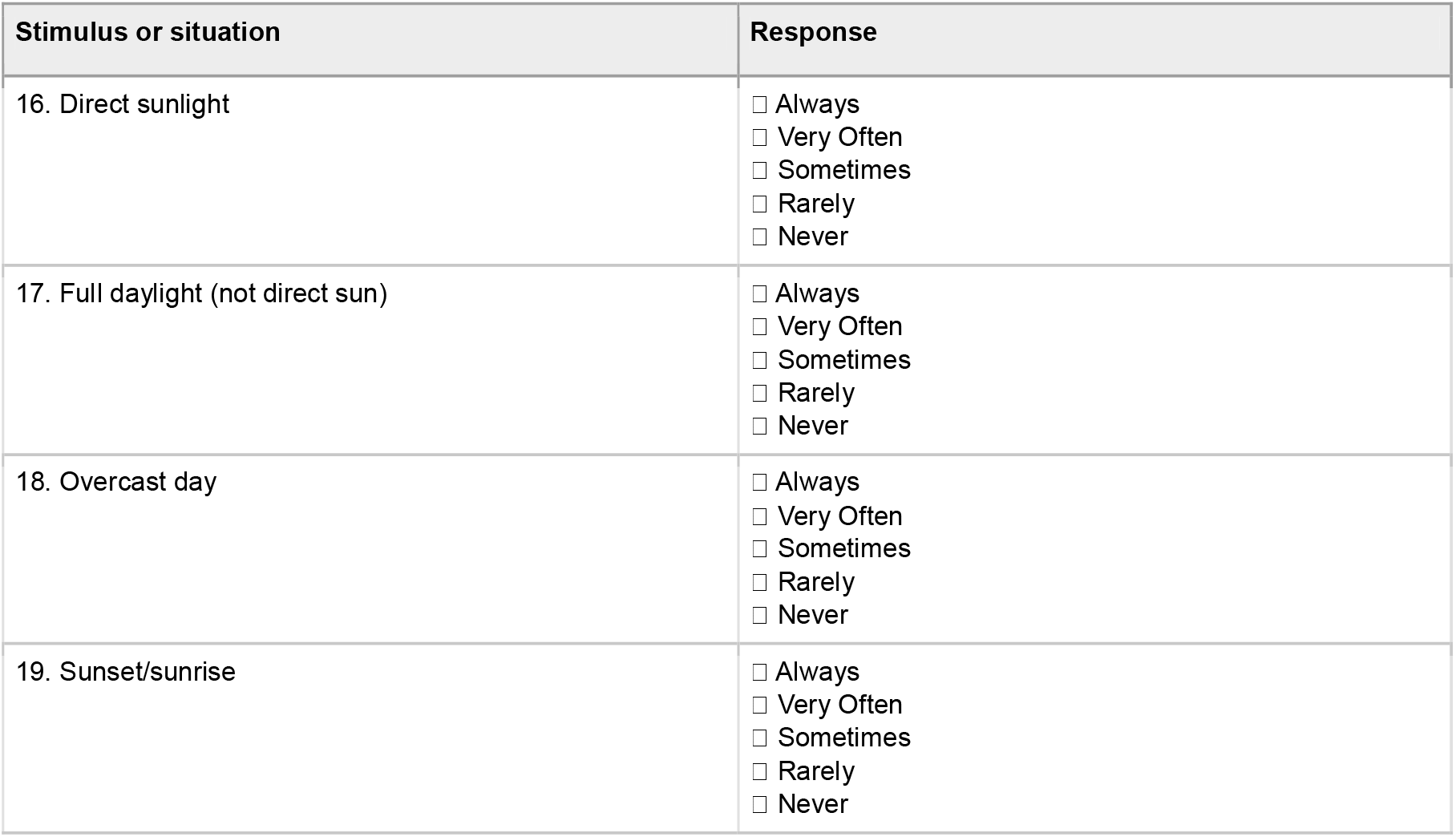
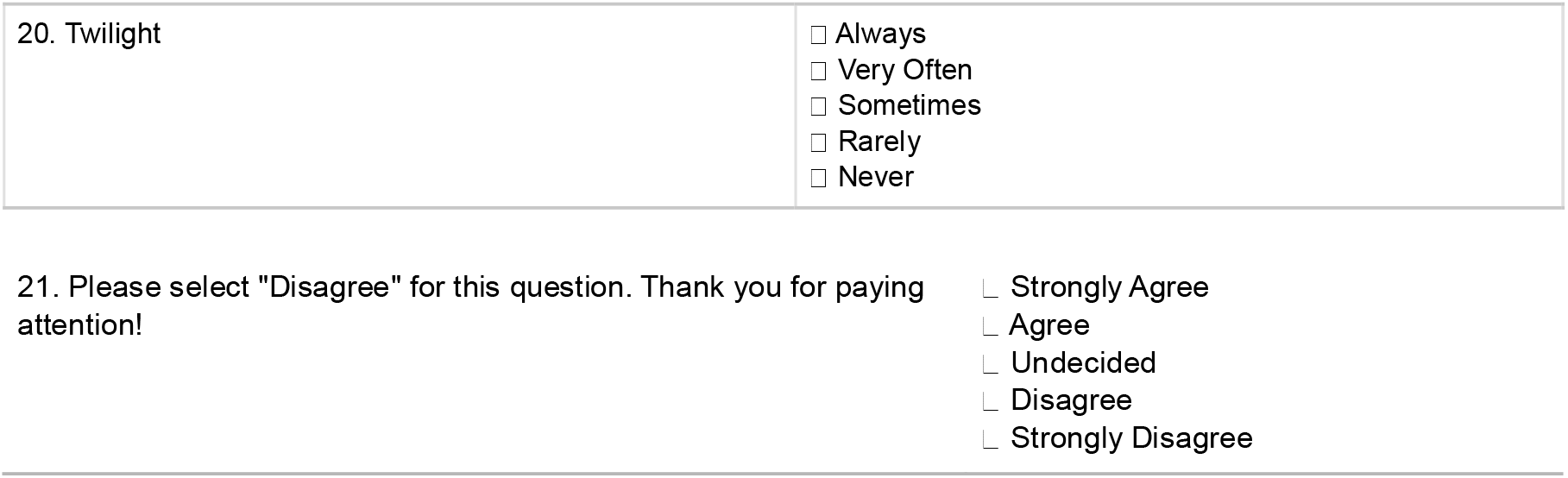

Sneezing in daylight Please select to what extent exposure to different levels of DAYLIGHT elicits a sneeze.
| Stimulus or situation | Response |
| --- | --- |
| 16. Direct sunlight | ☐ Always<br>☐ Very Often<br>☐ Sometimes<br>☐ Rarely<br>☐ Never |
| 17. Full daylight (not direct sun) | ☐ Always<br>☐ Very Often<br>☐ Sometimes<br>☐ Rarely<br>☐ Never |
| 18. Overcast day | ☐ Always<br>☐ Very Often<br>☐ Sometimes<br>☐ Rarely<br>☐ Never |
| 19. Sunset/sunrise | ☐ Always<br>☐ Very Often<br>☐ Sometimes<br>☐ Rarely<br>☐ Never |
| 20. Twilight | ☐ Always<br>☐ Very Often<br>☐ Sometimes<br>☐ Rarely<br>☐ Never |

21. Please select "Disagree" for this question. Thank you for paying attention!
☐ Strongly Agree ☐ Agree ☐ Undecided ☐ Disagree ☐ Strongly Disagree

## Sneezing in electric light

Please select to what extent exposure to different levels of ELECTRIC LIGHT elicits a sneeze.

| Stimulus or situation | Response |
| --- | --- |
| 22. Bright room illumination | ☐ Always<br>☐ Very Often<br>☐ Sometimes<br>☐ Rarely<br>☐ Never |
| 23. Moderate room illumination | ☐ Always<br>☐ Very Often<br>☐ Sometimes<br>☐ Rarely<br>☐ Never |
| 24. Dim room illumination | ☐ Always<br>☐ Very Often<br>☐ Sometimes<br>☐ Rarely<br>☐ Never |
| 25. Monitors, smartphone displays, TVs | ☐ Always<br>☐ Very Often<br>☐ Sometimes<br>☐ Rarely<br>☐ Never |
| 26. Direct view of light sources (e.g. lamps) | ☐ Always<br>☐ Very Often<br>☐ Sometimes<br>☐ Rarely<br>☐ Never |
| 27. Car headlights (at night) | ☐ Always<br>☐ Very Often<br>☐ Sometimes<br>☐ Rarely<br>☐ Never |
| 28. Other (please describe below) | ☐ Always<br>☐ Very Often<br>☐ Sometimes<br>☐ Rarely<br>☐ Never |

## Aspects of light making you sneeze

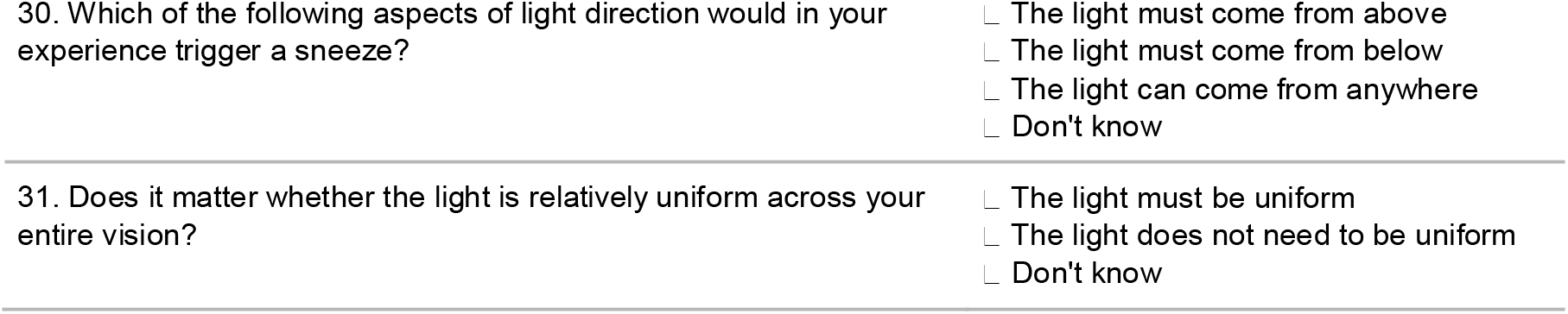

## Time of day

In this section, we would like to ask you whether there are any specific times during the day that you are more likely to sneeze due to BRIGHT LIGHT.

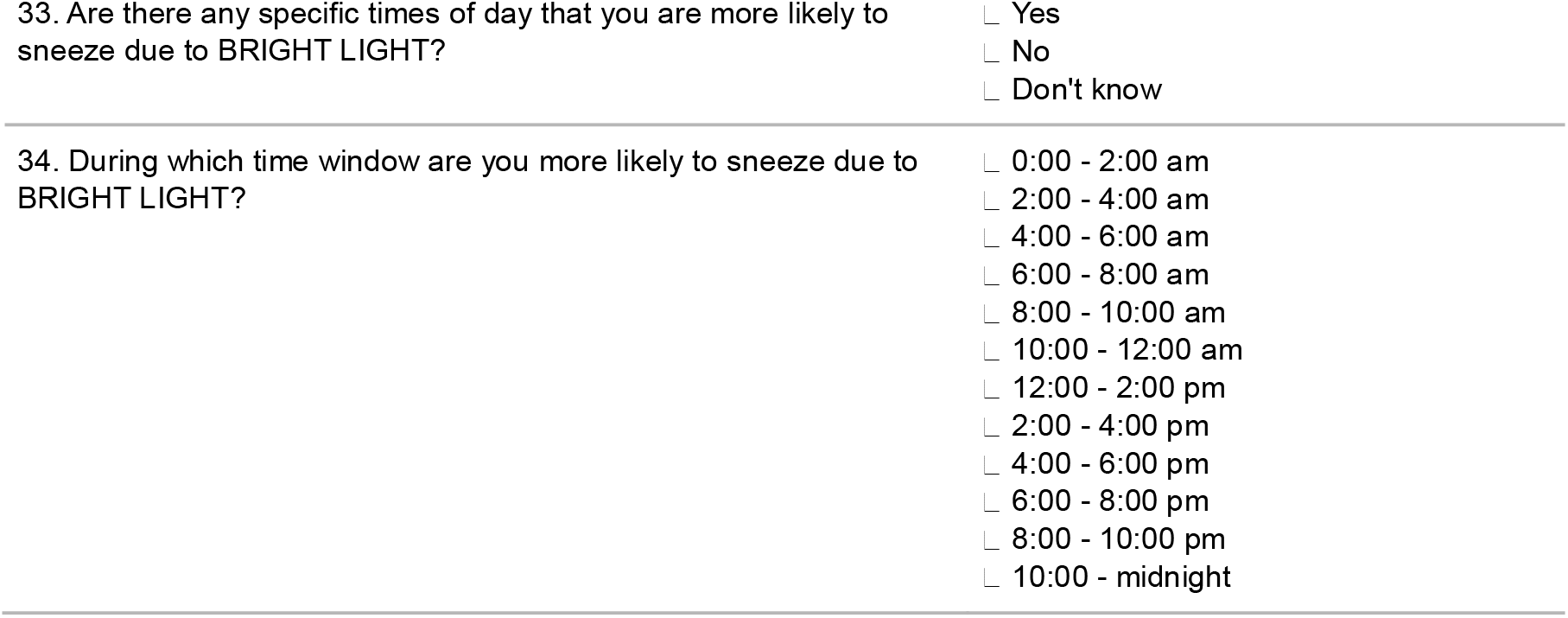

## Season

In this section, we would like to ask you whether there are any specific seasons during which you are more like to sneeze in response to bright light.

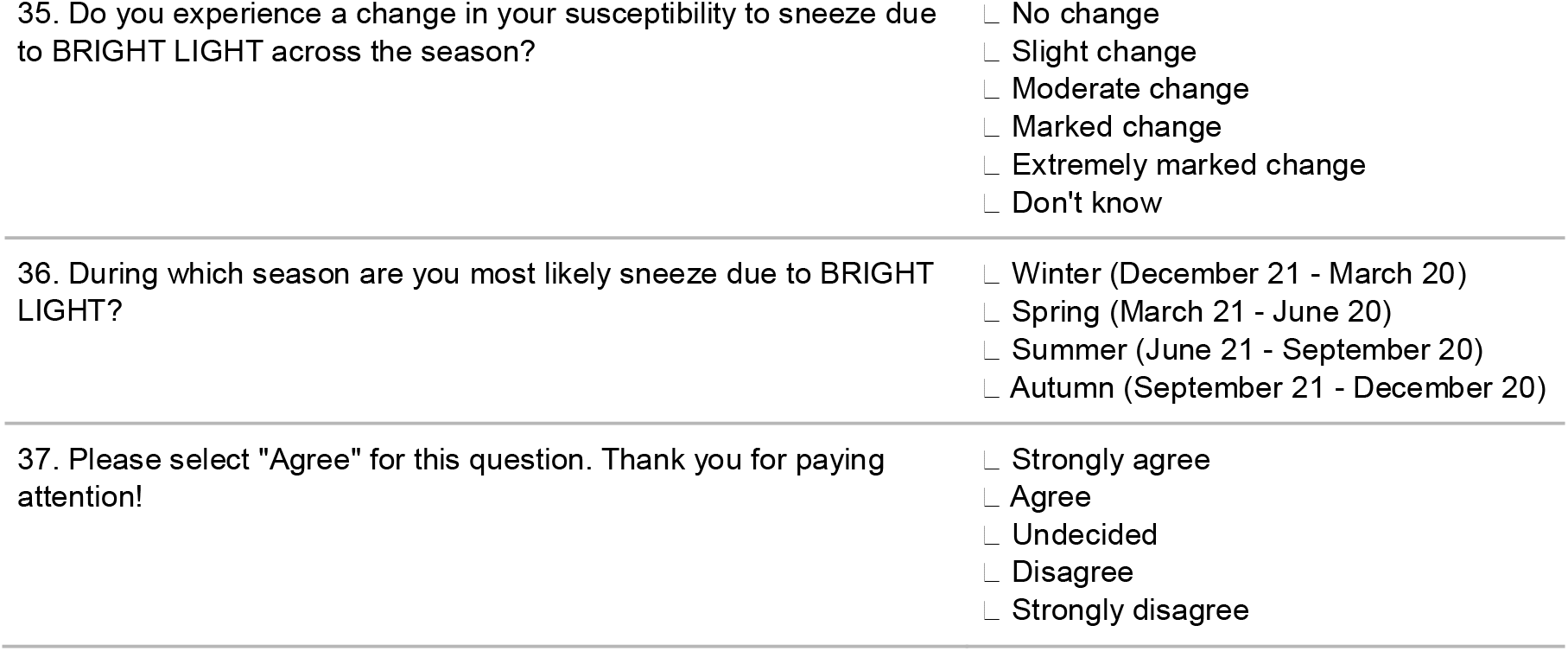

## Sneezing due to other stimuli

We would like to know about other stimuli or objects that make you sneeze. Please select to what extent other stimuli elicit a sneeze.

| Stimulus or situation | Response |
| --- | --- |
| 38. Inhaling cold air | ☐ Always<br>☐ Very Often<br>☐ Sometimes<br>☐ Rarely<br>☐ Never |
| 39. Inhaling dust | ☐ Always<br>☐ Very Often<br>☐ Sometimes<br>☐ Rarely<br>☐ Never |
| 40. Eating chocolate | ☐ Always<br>☐ Very Often<br>☐ Sometimes<br>☐ Rarely<br>☐ Never |
| 41. Eating mint or mint-flavoured products | ☐ Always<br>☐ Very Often<br>☐ Sometimes<br>☐ Rarely<br>☐ Never |
| 42. Smelling mint or mint-flavoured products | ☐ Always<br>☐ Very Often<br>☐ Sometimes<br>☐ Rarely<br>☐ Never |
| 43. Smelling pepper | ☐ Always<br>☐ Very Often<br>☐ Sometimes<br>☐ Rarely<br>☐ Never |
| 44. Eating spicy foods | ☐ Always<br>☐ Very Often<br>☐ Sometimes<br>☐ Rarely<br>☐ Never |
| 45. Drinking wine | ☐ Always<br>☐ Very Often<br>☐ Sometimes<br>☐ Rarely<br>☐ Never |
| 46. Smelling perfume and fragrances | ☐ Always<br>☐ Very Often<br>☐ Sometimes<br>☐ Rarely<br>☐ Never |
| 47. Smelling cleaning products | ☐ Always<br>☐ Very Often<br>☐ Sometimes<br>☐ Rarely<br>☐ Never |
| 48. Experiencing sexual arousal | ☐ Always<br>☐ Very Often<br>☐ Sometimes<br>☐ Rarely<br>☐ Never<br>☐ Don't know |
| 49. Experiencing strong emotions | ☐ Always<br>☐ Very Often<br>☐ Sometimes<br>☐ Rarely<br>☐ Never<br>☐ Don't know |
| 50. Plucking eyebrows | ☐ Always<br>☐ Very Often<br>☐ Sometimes<br>☐ Rarely<br>☐ Never<br>☐ Don't know |
| 51. Plucking nose hair | ☐ Always<br>☐ Very Often<br>☐ Sometimes<br>☐ Rarely<br>☐ Never<br>☐ Don't know |

## Some information about you

Finally, we would like to know a few things about you.

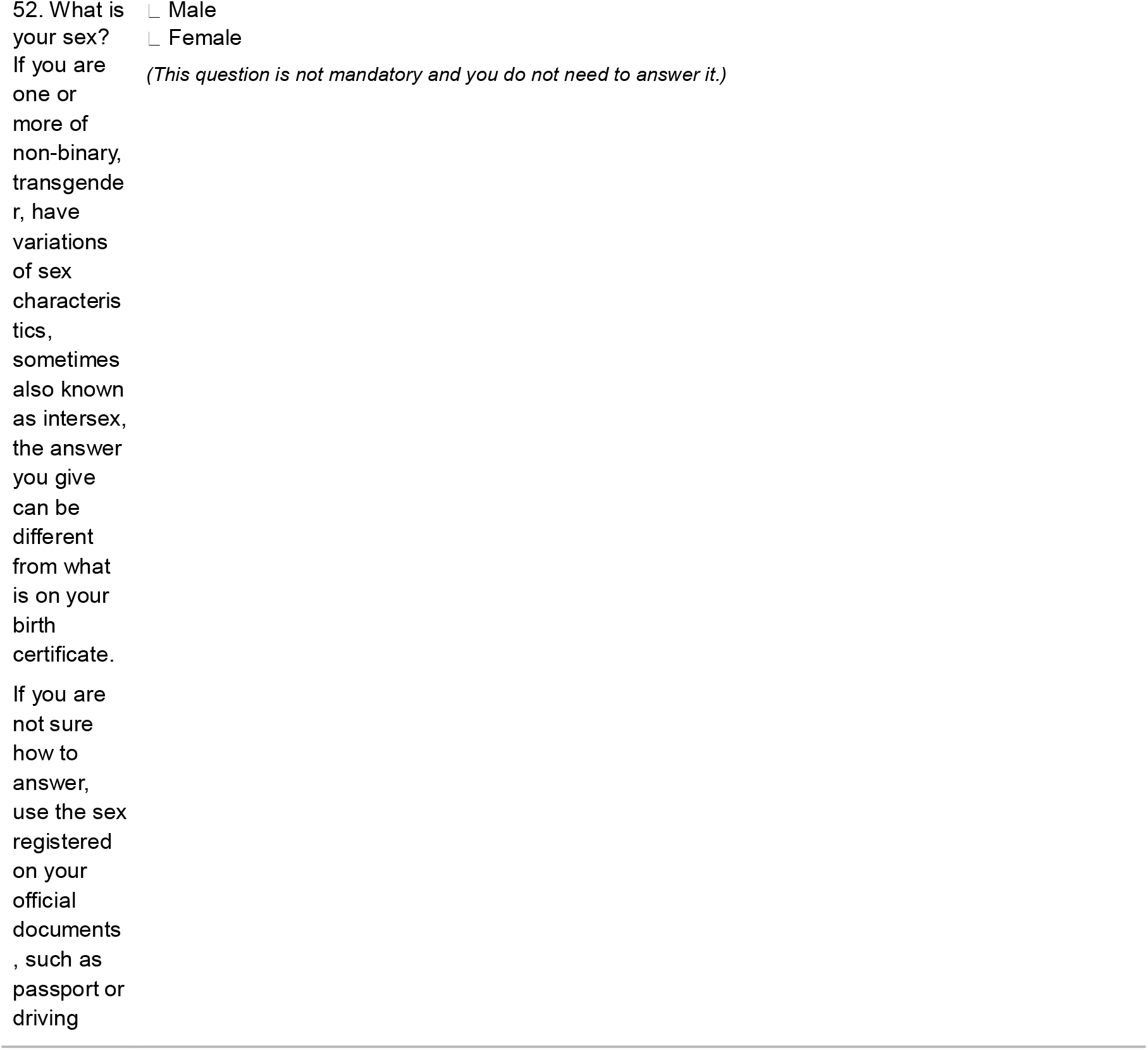
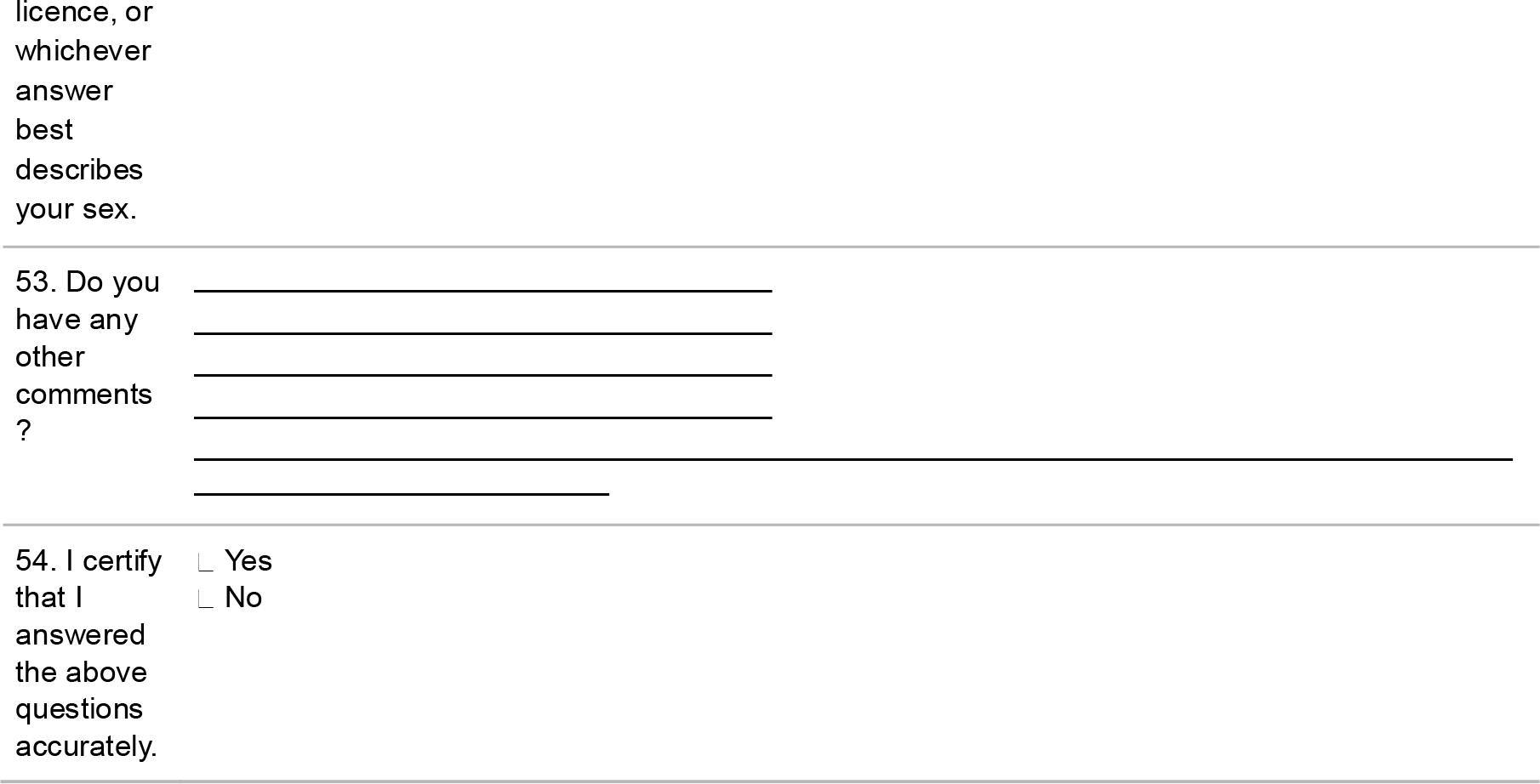

## Researcher note

Ancestry and ethnicity, relevant health status, and medication use are not included as questionnaire items here. Depending on the study aims, population, and analysis plan, researchers should consider collecting these data separately when and if applicable.

